# Hospitalization time and outcome in patients with Coronavirus Disease 2019 (COVID-19): analysis data from China

**DOI:** 10.1101/2020.04.11.20061465

**Authors:** Songshan Chai, Dongdong Xiao, Qikai Cheng, Shengzhi Huang, Yihao Wang, Jiajia Qian, Jiajing Wang, Junjun Li, Peng Fu, Qiangping Wang, Jing Rao, Hongyang Zhao, Fangcheng Zhang, Nanxiang Xiong

**Author notes:** Corresponding authors:Prof. Fangcheng Zhang, Department of Neurosurgery, Union Hospital, Jiefang Avenue No.1277, Wuhan 430022, China;, Prof. Nanxiang Xiong, Department of Neurosurgery, Union Hospital, Jiefang Avenue No.1277, Wuhan 430022, China. Chai Songshan, Dongdong Xiao, and Qikai Cheng contribute equally to this article.

## Abstract

**Objective:** The mean hospitalization time and outcome among patients with coronavirus disease 2019 (COVID-19) was estimated with the purpose of providing evidence for decision-making in medical institutions and governments in epidemic areas.

**Method:** The data of COVID-19 patients in china were collected from the websites of provincial and municipal health commissions. The mean hospitalization time and mortality in the mild or severe patients and the mean time from severe to mild illness were calculated by Gaussian mixture modeling.

**Results:** The mean hospitalization time among mild patients in Hubei province, other areas except Hubei province, and the national areas was 20.71± 9.30, 16.86 ± 8.24, and 19.34 ± 9.29 days, respectively. The mean transition time from severe to mild group in the above three areas were 15.00, 17.00, and 14.99 days, respectively. The death rate of mild and severe patients in Hubei province and the national areas were 1.10% and 18.14%, and 1.10% and 17.70%, respectively. Among those patients who died of COVID-19, the mean time from severe transition to death in Hubei province and the national areas was 6.22 ± 5.12 and 6.35 ± 5.27 days, respectively.

**Conclusion:** There were regional differences in the average length of stay between Hubei province and other regions, which may be related to different medical configurations. For those severe patients who died of COVID-19, the average time from hospitalization to death was about one week, and proper and effective treatments in the first week were critical.

## Introduction

The coronavirus disease 2019 (COVID-19) has become a major challenge to global health systems. Although the outbreak in China has been effectively controlled as yet, the COVID-19 is still spreading in other countries, posing a severe challenge to medical resources in all regions of the world.^1^ There have been many studies on the clinical manifestations, epidemic characteristics and treatment methods for infected patients, but the overall hospitalization time and outcome of patients with novel coronavirus pneumonia have not yet been systematically summarized.^2,3^ We deem that revealing the mean hospitalization of COVID-19 patients could provide valuable data to clinicians and local authorities that are bearing the brunt of the pandemic at present. From a political and economic perspective, our study can also provide the basis for resource integration in emergency circumstances. Here we attempt to make statistics on pneumonia data published daily by Chinese national and local governments since the outbreak of the epidemic in China. By using Gaussian mixture modeling, the informatic data analysis we obtained, indicators about the outcome of mild and severe patients were calculated.

## Material and methods

### Data sources

Data are based on information on outbreaks reported daily by provincial and municipal health commissions and compiled by the national health commission. Data obtained on a daily basis included: new confirmed cases, cumulative confirmed cases, new deaths on the same day, cumulative deaths, new discharge cases on the same day, and cumulative discharge cases. Because the number of COVID-19 patients in Hong Kong, Macao, and Taiwan is relatively small compared with the mainland region, cases in the three regions are not discussed.

### Diagnostic criteria (for China)

The patient’s diagnostic criteria were in accordance with the trial program 1 to 7 of Guidance for Corona Virus Disease 2019 issued by the national health commission.^4^ Besides, the China CDC organized national level training courses for medical staff in primary health agencies to ensure diagnostic accuracy. Concrete judgment standard is as follows:

1. Epidemiological history:
  1. Having a history of travel or residence in Wuhan and its surrounding areas or other communities with case reported within 14 days before the patient’s onset; or
  2. Having a contact history with patients (a positive results of nucleic acid test of 2019-nCoV) within 14 days before the patient’s onset; or
  3. Having a contact history with patients with fever or respiratory symptoms from Wuhan and its surrounding areas, or the communities with cases reported with 14 days before the patient’s onset; or
  4. Clustering occurrence of cases
2. Clinical manifestation:
  1. Fever and/or respiratory symptoms;
  2. Having the imaging features of pneumonia described above
  3. In the early stage, a normal or decreased total white blood cell count and a decreased lymphocyte count can be found

Sputum, pharyngeal swab or lower respiratory tract secretions, etc, were tested COVID-19 nucleic acid positive by real-time fluorescence RT-PCR; gene sequence of the virus showed high homology with the known COVID-19.

### Patient discharge standards (China)

According to Guidance for Corona Virus Disease 2019 issued by the national health commission, specific criteria are as follows:

1. With normal body temperature for more than 3 days;
2. With significantly recovered respiratory symptoms;
3. Lung imaging shows obvious absorption and recovery of acute exudative lesion;
4. With negative results of the nucleic acid tests of respiratory pathogens for consecutive two times (sampling interval at least 1 day)

### clinical classification of novel coronavirus pneumonia

According to the Guidance for Corona Virus Disease 2019, patients were classified as mild, general, severe and critical according to the severity of their clinical presentation. During data statistics, we classified mild and general patients as the mild group, severe and critical patients as the severe group. During our study, patients deteriorated and transformed to severe condition and critical cases were ultimately classified as the severe group. Severe patients have two outcomes, one is cured and discharged, the other is death. Then, we calculated the time from admission to discharge and to death respectively for patients in severe group.

### Data processing

#### Model definition

The number of daily discharges of COVID-19 cases in each province can be divided into the sum of the number of discharges of patients with severe condition and the number of discharges of patients with mild condition. By observing the daily distribution of discharged patients in major regions, we found that the distribution showed a multi-peak situation. So we assume that the distribution of the number of new cases of COVID-19 discharged per day in each province conforms to a Gaussian mixed distribution, in which the probability of discharge from severe patients after Hospitalization *j* days and the discharge probability of mild patients after hospitalization *j* days are fitted with normal distributions. In addition, we assume that the severe disease rate *r*, the mild disease mortality rate *R*_1_ and the severe disease mortality rate *R*_2_ follow normal distribution respectively, that is, the number of severe patients in the newly added cases on the day can be estimated by *xi·r*, and *x*_*i*_·(1 − *r*) can evaluate the number of mild patients in the newly added cases on the same day (*x*_*i*_ represents the number of newly added cases on the same day *i*).

Specifically, the estimated number of discharged patients on day *n* is equal to the sum of the estimated number of discharged patients on day 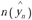 and the estimated number of discharged patients on day 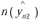, that is:

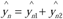

Among them, 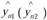 can be calculated by the cumulative sum of the number of patients with discharge who were admitted to the hospital before day *n* after treatment for j days (*j* = *n* − *i*), Which satisfies the following equation: 

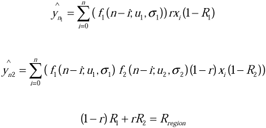

*f*_1_ represents the discharge probability of patients with mild disease and *f*_2_ represents the probability of severe patients converting to mild one. Therefore, the discharged severe patients were first converted to mild patients and then discharged, and the discharged mild patients were discharged directly after recovery. Mild mortality *R*_1_ and severe mortality *R*_2_ meet the constraints of mortality *R*_*region*_ in the region to be estimated, where *R*_*region*_ can be obtained by collecting data. Severe disease rate *r* was a constant of 0.195551694 in Hubei province at the time of correlation fitting, and was set as the parameter to be estimated in the interval in other regions due to the unknown *r*. (here we idealize the assumption that discharged patients do not experience the repeated cycle from mild to severe to mild).

We explain the model with the table 1 and figure 1. The table 1 shows the composition of the number of patients discharged from the hospital on the day *n*. The first line indicates the time of admission, *j*. Lines 2 and 3 indicate the number of discharged patients after *j* day of hospitalization for mild and critical patients, respectively. As shown in Figure 1,The bottom blue and red dashed interval curves show the normal distribution curve of the number of discharged mild and critical patients in *day*_0_, respectively. The green and orange dotted dashed interval curves in the middle indicate the normal distribution curve of the number of discharged mild and severe patients in *day*_1_, respectively. The top black curve represents the distribution of the sum of the number of mild and critical patients discharged in *day*_0_ and *day*_1_.

**Table 1.**
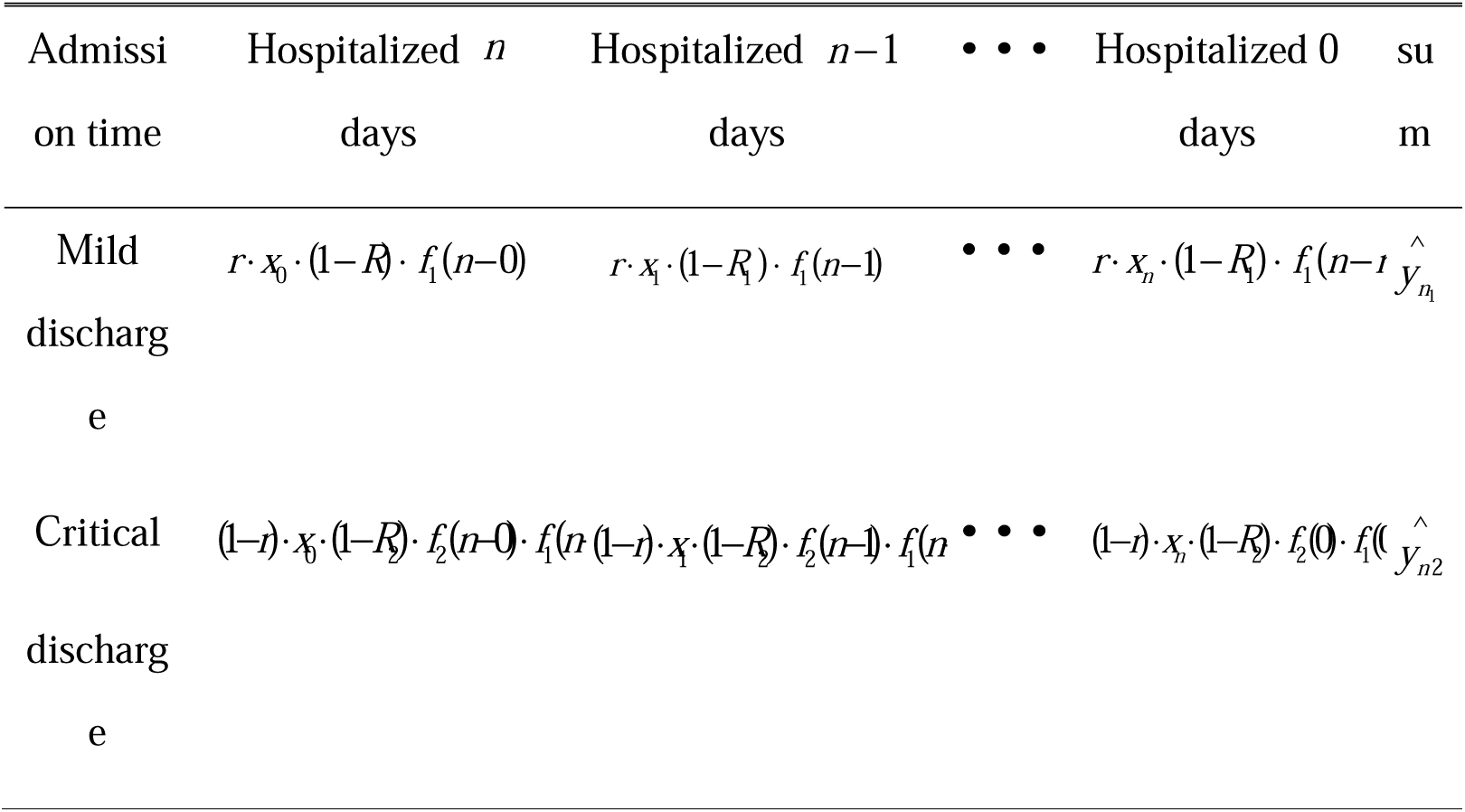
the composition of the number of patients discharged from the hospital on the day *n*

**Figure 1:**
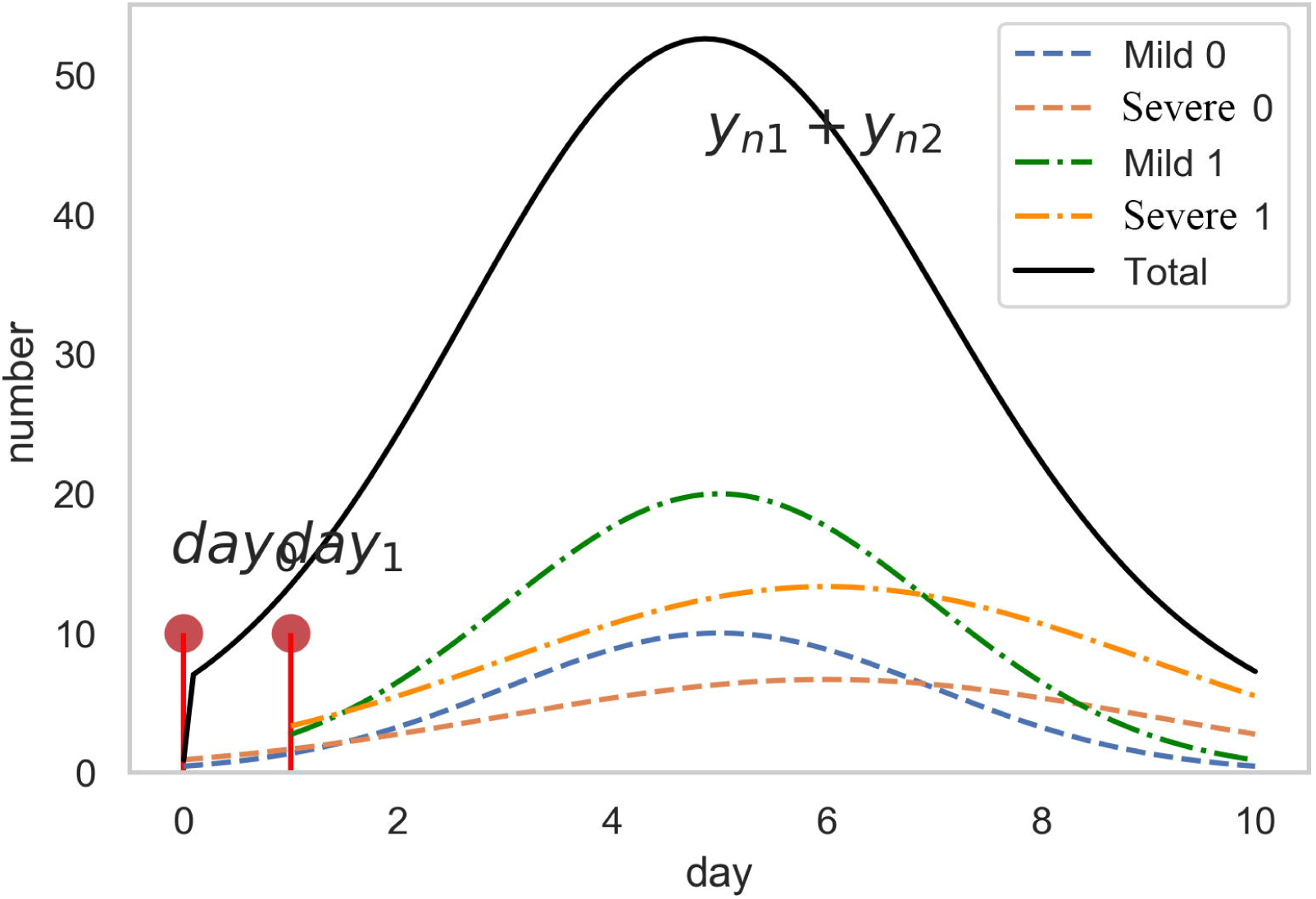
A defined curve simulating the number of daily discharges or deaths among mild and severe patients the bottom blue and red dashed interval curves show the normal distribution curve of the number of discharged mild and critical patients in *day*_0_, respectively. The green and orange dotted dashed interval curves in the middle indicate the normal distribution curve of the number of discharged mild and severe patients in *day*_1_, respectively. The top black curve represents the distribution of the sum of the number of mild and critical patients discharged in *day*_0_ and *day*_1_.

### Model definition for death patients

Specifically, the estimated number of deaths 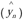 on the day(*n*) is equal to the sum of the number of deaths 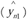 of mild patients on the same day and the number of deaths 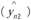 of severely ill patients on that day.

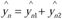

In which, 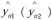 can be calculated from the accumulative sum of the total number of deaths of mild (severe) patients admitted to hospital before the days *n* on the day (*j*) of treatment (*j* = *n* − *i*). It satisfies the following equation:

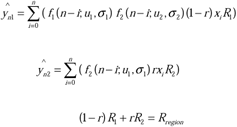

Therefore, dead mild patients first become severe and then die (here we idealize the hypothesis that the dying patients do not experience the repeated cycle from mild to severe to mild).

Therefore, the average time for mild patients from admission to death is *u*_1_ + *u*_2_, and the average time severe patients is *u*_2_.

### Quadratic cost function

We take the squared residuals as the optimization goal, so we have the following cost function

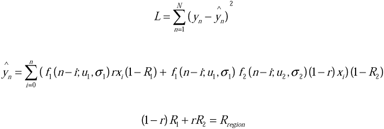

In the above formula, *N* represents the total number of observation days. *y*_*n*_ indicates the actual number of patients discharged on nth day, 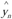 represents the estimated number of discharged patients on the nth day of the model. Where 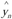 is fitted using a Gaussian mixture model, *f*_1_ and *f*_2_ represent normal distribution functions with mean *u*_1_ (*u*_2_) and variance *σ*_1_ (*σ* _2_), respectively. *r* is the critical rate. *x*_*i*_ is the number of new patients on ith day. 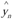 means that the number of patients discharged on the nth day is equal to the number of patients discharged on the day before the day *n*.

We use the Newton iteration method to solve the optimal solution of the cost function 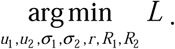.

### Newton iterative algorithm

In the following, we first introduce the operation process of Newton’s iteration method and then provide the partial derivatives required by Newton’s iteration method.

First, we record *θ* as the estimated parameter. When the critical rate *r* is constant, *θ* equals (*u*_1_,*u*_1_,*σ*_1_, *σ* _2_, *r*), and when the critical rate *r* is an optimized parameter, *θ* equals (*u*_1_, *u*_1_, *σ*_1_, *σ* _2_, *r, R*_1_, *R*_2_).

1. Given an initial value of *θ, θ*_0_.
2. 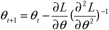
3. if ‖*θ*_*t* +1_ − *θ*_*t*_ ‖< *ε*, then stop iterating. Otherwise, skip to step (2) to update parameters*θ*.

In the numerical experiment part, we use the curve-fit function in the scipy. Optimize package in Python to help implement the Newton algorithm iterative process.

Because the Newton iteration method used in this paper is sensitive to initial values, we expect to determine the effective initial value of the Newton iteration method by the grid method to ensure better curve fitting. In the grid search process, after adjusting the search space of the parameter *θ* to be estimated multiple times, we select the parameter estimation value with the best curve fitting effect in the grid range selection method as the initial value of the Newton iteration method. For the sake of simplicity, here we omit the experimental process of grid range selection and the curve fitting results of the grid method, and only show the curve fitting results obtained by the Newton iteration method.

## Results

Using the gauss-type function,we calculated the curve fitting results of Hubei province, provinces together except Hubei, whole of China that obtained by the Newton iteration method. Through the moving average method, the red curve in the above figure **(Figure1)** is smoothed to a certain degree, and the curve fitting effect we proposed is also better. Eventually, the simulated data and the actual situation fit well. Furthermore, the mean hospitalization, the time window of patient progression and deterioration were calculated. The detailed fitting results are as follows.

### 1. Curve Fitting Results in Hubei Province

During the curve fitting experiment, we also tried to set the COVID-19 critical rate *r* in Hubei Province as an optimizable parameter in the interval [0.1,0.25]. However, since the patient data in Hubei Province are sufficient, the effect of *r* is small. So we do not give a fitting figure here. The severe disease rate in Hubei province was set as 0.195551694.

Hubei has the highest number of cases with 51,796. Curve fitting results (**Figure2A**) showed that the transition time from severe to mild is about 15 days and for mild the time to discharge is 20.71±9.3 days. We can see that the mean hospitalization time of mild patients fluctuates greatly. Accordingly, the mean hospitalization for severe patients was 35.71 days.

**Figure 2.**
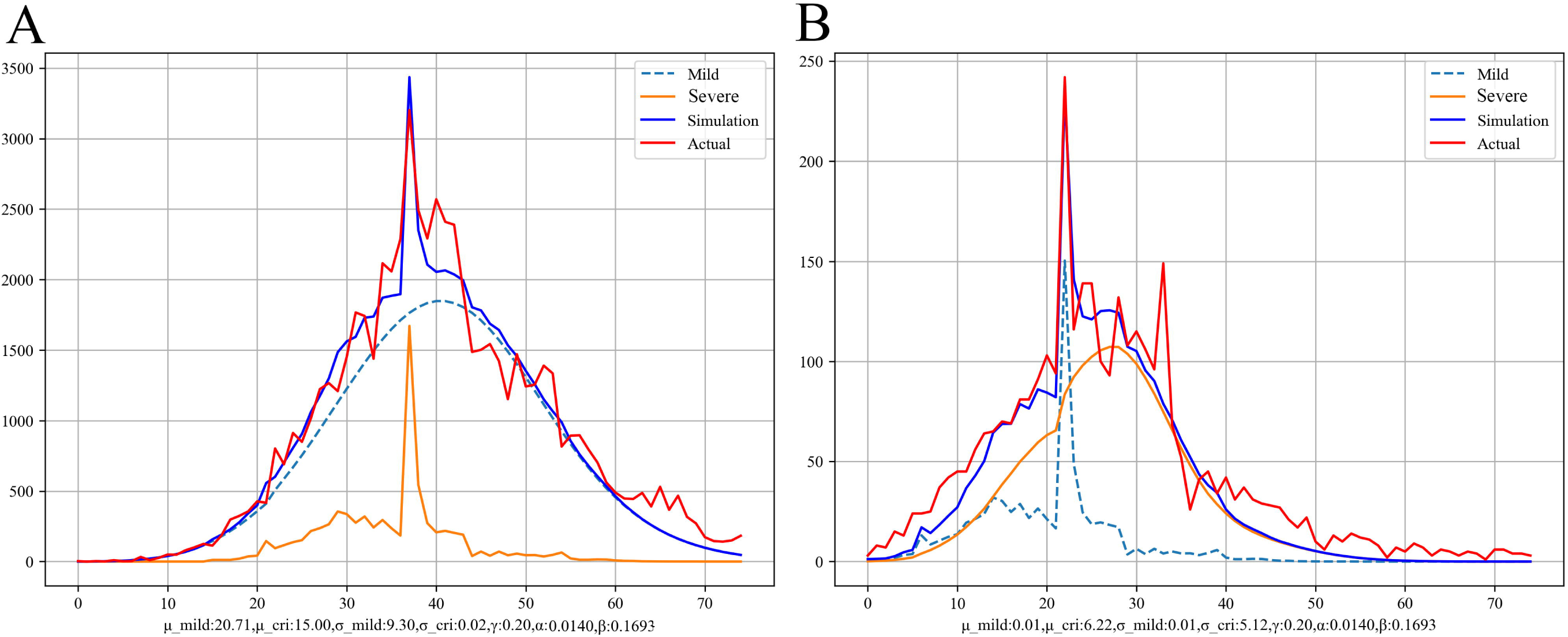
Curve fitting graphs of daily discharge numbers (A) and daily death (B) numbers in Hubei province. The light blue dotted line and orange solid line curve represent the results of mild and severe patients, respectively. The dark blue and red solid lines represent the simulated and actual results of all patients.

**Figure2B** reflects the fitting curve of the time from admission to death of COVID-19 infected patients in Hubei province. For the patients who died, they were divided into mild patients and severe patients at the time of admission. According to the curve fitting result, mild cases that eventually die tend to deteriorate rapidly and the time is within one day for most patients in this group. This indicates that patients who showed mild symptoms on admission and eventually die have a rapidly deteriorating condition, which requires clinicians to prepare such patients for rescue in advance. The time from admission to death for severe patients is ranges from 6.22±5.12 days. Overall, the death rate of mild cases was 0.0140 and that of severe cases was 0.1693.

### 2. Curve Fitting Results At The National Level

Research at the national level showed that time required from severe to mild is similar to Hubei province and from admission to cure and discharge of mild cases the time is about 19.34±9.29 days. So, the mean hospitalization at the national level for severe patients was 28.63 days, the time required from severe to mild add time from mild to discharge. we set the critical rate *r* in other provinces as an optimized parameter range from 0.1 to 0.25.(**Figure3A**) Death data of the national level was showed in **Figure3B**. Time from admission to death for severe patients was 6.35±5.27 days.

**Figure 3.**
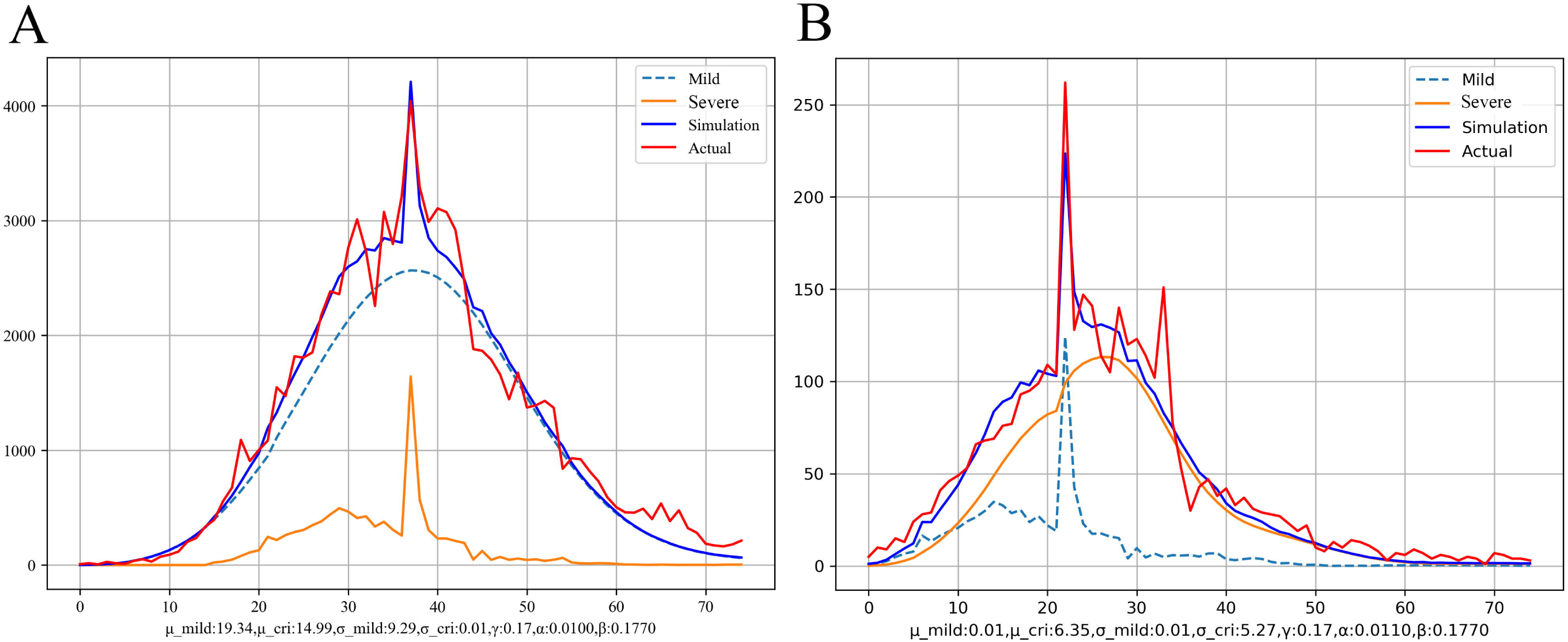
Curve fitting graphs of daily discharge numbers (A) and daily death (B) numbers in the national areas. The light blue dotted line and orange solid line curve represent the results of mild and severe patients, respectively. The dark blue and red solid lines represent the simulated and actual results of all patients.

### 3. Curve Fitting Results in Other Areas Except Hubei Province

Compared with Hubei province, other provinces have fewer cases. For the curving fitting, death data in other provinces is too little, this leads to great deviation in the simulative curve and we can only simulate the discharge time fitting curve (**Figure4**). We deem that the mean hospitalization of mild disease is 16.86±8.24 days, which was shorter than that of hubei province. The time required for a severely infected patient to become mild is 17.00 days on average. The corresponding mild and severe death rates were 0 and 0.0633 respectively, which is obviously lower than the level of Hubei province. The mean hospitalization time was 33.86 days and the *r* was 0.17.

**Figure 4.**
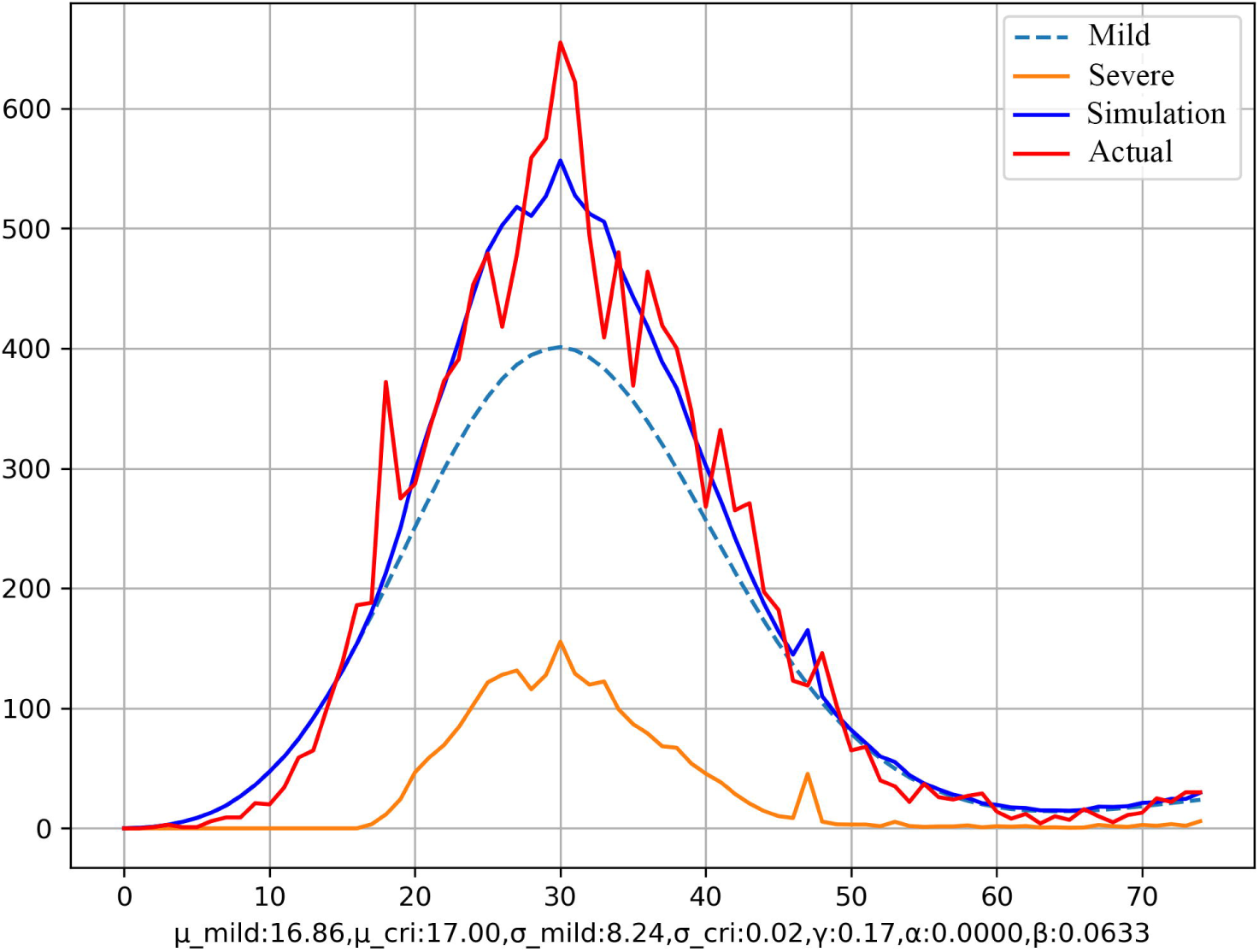
Curve fitting graphs of daily discharge numbers in other areas except Hubei province. The light blue dotted line and orange solid line curve represent the results of mild and severepatients, respectively. The dark blue and red solid lines represent the simulated and actual results of all patients.

## DISCUSSION

COVID-19 is an infectious disease that appears at the end of 2019. It has a strong ability to spread and has a high fatality rate in severe patients.^1-3^ At present, the control of the COVID-19 epidemic and the treatment of infected patients is a huge challenge facing humans. Isolating infected people from uninfected people is the most important measure to control the spread of this infectious disease. At the same time, patients with this pneumonia also need to be actively treated. In China, all diagnosed patients will be isolated and treated by designated hospitals or mobile cabin hospitals.^5-7^ Basically, the length of hospitalization is the time to cure or die. Because of the great difference in treatment and prognosis between mild and severe patients, it is necessary to calculate and discuss the mean hospitalization time for mild and severe patients separately.

In the mild patients, after the onset of mild illness, ground-glass-like shadows appear in the lungs, and the inflammation of the lungs generally peaks at 9-13 days.^8^ If the disease does not continue to progress, it will improve on its own in 2 weeks or even more.^9,10^ The cured patients could be discharged without waiting for imaging to be completely normal. For the treatment of mild patients, during the peak of the epidemic in China, many stadiums or large factories were converted into mobile cabin hospitals, where mild patients were admitted and observed medically. The mobile cabin hospitals can provide basic medical support. The reason why mild patients were admitted to the mobile cabin hospitals is that they can be detected by medical staff in time and transferred to a nearby designated hospital for treatment once getting worse. The problem of the isolation and treatment of a large number of mild patients was quickly and economically solved by building the mobile cabin hospitals. Our simulation results showed that the hospital stay of mild patients is different in different regions; the hospitalization time among mild patients in the national areas was range from 9.3 to 30.0 days. It is slightly longer in Hubei province than in other regions. The mean hospitalization time required for mild patients could be used as a reference for the number of medical supplies and medical personnel to be prepared.

We divide the improvement process of severe patients into two stages. The first step is from severe to mild illness, and the second step is from mild illness to cured and discharged. According to the results, in Hubei province and the national areas, the mean transition time from severe to mild illness was both about 15 days; the time from mild illness to discharge is 20.71 days, Therefore, the mean hospitalization time among severe patients who were ultimately cured was 35.71(20.71 add 15.00) days. This similar result could be explained by the fact that the majority of domestic COVID-19 cases exist in Hubei. From our statistics, in Hubei province, the death rate in all patients was 4.51%, and the death rate in severe patients was 18.14%. In fact, we also found that the daily death rate showed a significant downward trend with time (data not shown), especially in the later stage of the epidemic, where the treatment experience of critically ill patients has increased and the treatment methods have been diversified. Our study also showed that the average time for severe patients in Hubei province and the national areas from admission to death was 6.22 and 6.35 days, respectively. Therefore, the first week after admission is critical to manage severe patients. We found that those mild patients who eventually died of COVID-19 experienced a transition from mild to severe illness in a short time after admission (within 1 day). These rapidly deteriorating cases are consistent with previous reports.^11^ We could infer that these rapidly deteriorating cases may have experienced an inflammatory storm inducing a very high risk of death.

This study is the first to calculate the average length of hospitalization of patients with COVID-19 based on big data and Gaussian mixture modeling. Although not all patients have been discharged in China, Gaussian mixture modeling can still be used appropriately to calculate the length of stay of patients with mild and severe diseases. All our simulation results are based on the data as of April 4th, 2020, although it may take some time for all the remaining patients who are still in the hospital to have improved discharge or worsened death events, this has little effect on our results. At present,due to the epidemic in many countries is still spreading, the inflection point of the epidemic is far from coming, and many patients are still in the treatment stage. Therefore, the time course of patients with COVID-19 provided is an important reference for countries that are preventing and controlling the epidemic to estimate medical supplies and medical manpower required. We believe that our analysis data from China can provide references for other countries which are fighting the COVID-19 epidemic.

## Data Availability

The raw/processed data required to reproduce these findings cannot be shared at this time as the data also forms part of an ongoing study.

## Acknowledgement

This work was supported by Hubei Province Technological Innovation Special Soft Science Research Project (2019ADC034) to Xiong NX.

